# Time-to-event assessment for the discovery of the proper prognostic value of clinical biomarkers optimized for COVID-19

**DOI:** 10.1101/2021.07.09.21260262

**Authors:** José Raniery Ferreira, Victor Henrique Alves Ribeiro, Marcelo Cossetin, Marcus Vinícius Mazega Figueredo, Carolina Queiroz Cardoso, Bernardo Montesanti Almeida

## Abstract

In the early days of the pandemic, clinical biomarkers for COVID -19 have been investigated to predict patient mortality. A decision tree has been proposed previously comprising three variables, i.e., lactic dehydrogenase (LDH), high-sensitivity C-reactive protein (CRP), and lymphocyte percentage, with more than 90% accuracy in a public cohort. In this work, we highlighted the importance of the cohort made publicly available and complemented the findings by incorporating further evaluation. Results confirmed poor short-term prognosis to abnormal levels of some laboratorial indicators, such as LDH, CRP, lymphocytes, interleukin-6, and procalcitonin. In addition, our findings provide insights into COVID-19 research, such as key levels of fibrin degradation products, which are directly associated with the Dimerized plasmin fragment D and could indicate active coagulation and thrombosis. Still, we highlight here the prognostic value of interleukin-6, a cytokine that induces inflammatory response and may serve as a predictive biomarker.

## Main Text

In the early days of the pandemic, Dr. Yan and colleagues investigated clinical blood-originated biomarkers for COVID-19 to predict patient mortality [1]. The authors proposed a straightforward decision tree with three variables, i.e., lactic dehydrogenase (LDH), high-sensitivity C-reactive protein (hs-CRP), and lymphocyte percentage. They stated to have obtained high performance on an independent testing set with more than 90% accuracy. In this letter, we highlight the importance of their cohort made publicly available, especially at the beginning of the pandemic where open data were still limited, and complement the author’s findings by incorporating further evaluation.

Although it is an interesting approach, Dr. Yan and colleagues considered the problem as a classification task, which may not be the most proper way of dealing with continuous time-to-event data like survival [2-4]. As it is vastly known, machine-learning-based assessment is pruned to over-optimistic testing results using small sampling for training. In addition, it has been shown that their proposed model has limited performance on independent external datasets [5-7]. These two limitations are possibly due to model overfitting.

Therefore, in this study, we performed time-to-event analyses using the author’s original training dataset to find a proper predictive potential for the biomarkers investigated. Our evaluation aimed to optimize the clinical variables modeled in the previously developed decision tree and to discover other blood biomarkers with prognostic value for COVID-19. Opposing the original strategy [1], we also focused on identifying biomarkers for different sub-populations, according to the patient age and time of hospitalization.

We split the original training dataset into discovery and validation sets to perform a robust assessment and validate the results. The thresholds identified in the discovery set for risk stratification were then applied in the validation set to confirm the statistical significance and further performance. High and low-risk groups of patients were split according to the variable’s median value used as threshold identified in the discovery set [3, 4]. The log-rank test assessed the statistical difference between Kaplan-Meier curves and Cox proportional hazards regression models from stratified groups. The survival v3.2.3 and survminer v0.4.7 packages from R v4.1.0 performed the time-to-event analyses, with p < 0.05 being considered statistically significant.

The original training dataset comprised a few demographics data, i.e., patients’ age (varying from 18 to 95, averaging 58.8 ± 16.5 years old) and sex (224 men and 151 women), along with the results of 74 blood tests in different hospitalization times. As expected, the older the patient is, the worst is the prognosis [8, 9]; the threshold of 62 years obtained significant difference on survival curves (Figure 1.a).

**Figure 1.**
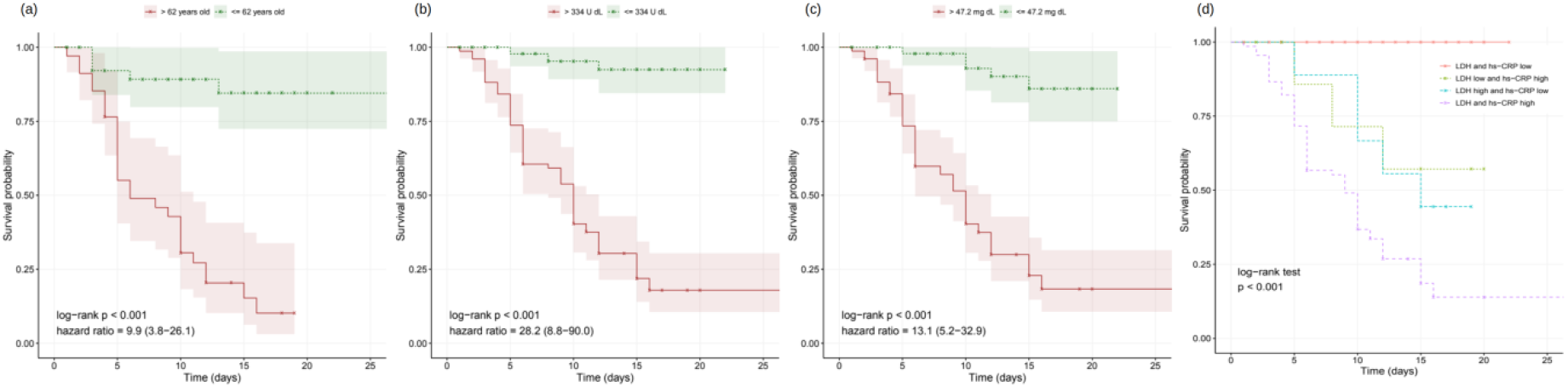
Kaplan-Meier curves of the clinical biomarkers of (a) age, (b) lactic dehydrogenase (LDH), (c) high-sensitivity C-reactive protein (hs-CRP), (d) LDH combined with hs-CRP.

The overall assessment disregarding patient age and hospitalization timing found predictive value on the validation set in 53 variables, including LDH (Figure 1.b), hs-CRP (Figure 1.c), and lymphocyte percentage. Besides those three, other biomarkers yielded relevant information on COVID-19 prognostication (Table 1). For instance, high-risk groups stratified by fibrin degradation products presented 97% likelihood of death and a hazard ratio (HR) of 4.26 (95% confidence interval (CI): 1.88-9.64); and elevated interleukin-6 (IL-6) associated with 65% likelihood of death and HR of 18.20 (CI: 2.42-136.54).

**Table 1.**
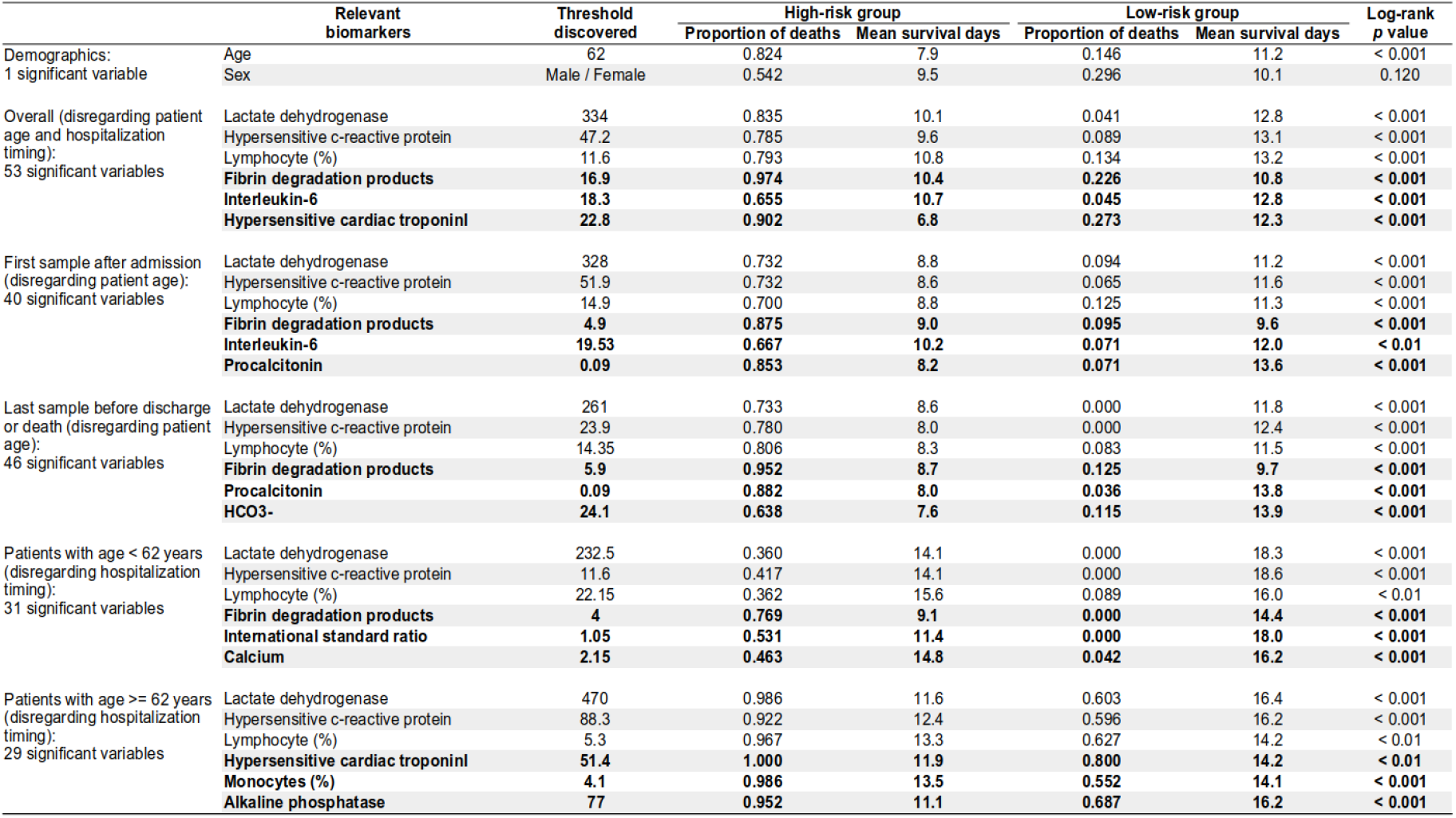
Discovered biomarkers according to the patient age and hospitalization time.

Furthermore, LDH and hs-CRP combined presented complementary predictive potential in multivariate assessment (Figure 1.d). With both biomarkers values elevated, patients showed a likelihood of death of 87%, mean overall survival time of 9.5 days, and HRs of 8.19 (CI: 2.27-29.52) and 3.90 (CI: 1.41-10.72). Conversely, when either LDH or hs-CRP yielded low value, potentially indicating lower risk, the age determined the worse prognosis in the multivariate signature (p < 0.001), resulting in a likelihood of death of 72% and HR of 7.01 (CI: 3.10-15.84) for the elderly patients.

Results confirmed poor short-term prognosis to abnormal levels of some laboratorial indicators, such as LDH [1, 9-11], CRP [1, 8-11], lymphocytes [1, 8, 10], IL-6 [12], and procalcitonin [11]. These findings could also provide insights into COVID-19 research, such as key levels of fibrin degradation products, which are directly associated with the Dimerized plasmin fragment D and could indicate active coagulation and thrombosis [9, 11]. Moreover, those biomarkers could identify the patient’s rapid worsening at early stages, when treatment is more likely to be successful, or before the clinical condition deteriorates at a critical time, avoiding later complications, like pulmonary embolism [13].

Dr. Yan and colleagues had already mentioned that lymphocytes may serve as a potential therapeutic target [1]. Still, we highlight here the role of IL-6, a cytokine that induces inflammatory response and has prognostic value. Although IL-6 blockade is not the standard strategy for COVID-19 treatment yet, interleukin-6 remains the best available biomarker for severity assessment and still holds great potential for targeted therapy [12, 14, 15].

In this work, we have identified relevant blood biomarkers that could be a mainstay for the clinical evaluation of COVID-19. In addition, these biomarkers could bring significant benefits to the COVID-19 clinical routine as they are fully available in medical practice, enabling the triage of higher-risk patients for quick treatment decision-making. Moreover, they correlated with short-term outcomes and could support the management of the disease with early interventions, ultimately leading to better endpoints such as decreased deterioration and mortality.

In future works, it would be essential to investigate long -term prognostication as morbidity may not always be seen in the acute stage of the disease. Furthermore, we highlight the importance of other research groups to further assess the biomarkers according to their population specificity [5-7]. Finally, a prospective evaluation of the biomarkers as the primary objective is necessary for robustness purposes and to confirm the findings.

## Data Availability

All data used in this work for experiments are publicly available.

## Notes

### Competing Interest Statement

The authors have declared no competing interest.

### Funding Statement

No external funding has been received.

### Author Declarations

No IRB approval was needed as the data used is publicly available.

